# Modelling the impact of Plasma Therapy and Immunotherapy for Recovery of COVID-19 Infected Individuals

**DOI:** 10.1101/2020.05.23.20110973

**Authors:** Nita H. Shah, Ankush H. Suthar, Ekta N. Jayswal, Nehal Shukla, Jagdish Shukla

## Abstract

Since the first case of COVID-19 was detected in Wuhan, China in December 2019, COVID-19 has become a pandemic causing a global economic and public health emergency. There is no known treatment or vaccine available for COVID-19 to date. Immunotherapy and plasma therapy has been used with satisfactory efficacy over the past two decades in many viral infections like SARS (Systemic Acute Respiratory Syndrome), MERS (Middle East Respiratory Syndrome), and H1N1. Limited data from China show clinical benefit, radiological resolution, reduction in viral loads, and improved survival. Our aim is to create a mathematical model for COVID-19 transmission and then apply various control parameters to see their effects on recovery from COVID-19 disease. We have formulated a system of non-linear ordinary differential equations, calculated basic reproduction R_0_, and applied five different controls (self-isolation, quarantine, herd immunity, immunotherapy, plasma therapy) to test the effectiveness of control strategy. Control optimality was checked by Lagrangian functions. Numerical simulations and bifurcation analyses were carried out. The study concludes that the COVID-19 outbreak can be controlled up to a significant level three weeks after applying all the control strategies together. These strategies lead to a reduction in hospitalization and a rise in recovery from infection. Immunotherapy is highly effective initially in hospitalized infected individuals however better results were seen in the long term with plasma therapy.

## 1. Introduction

The first case of unknown pneumonia was detected in Wuhan China in December 2019 and was later identified as COVID-19 spread by SARS COV-2 (severe acute respiratory syndrome coronavirus 2)(Cohen and Normile 2020). COVID-19 was declared as pandemic on 11th March by WHO. It has spread to 210 countries and as per worldometer (Worldometer 2020) and as of 25th April 2020 worldwide total cases are around 2,915,365 and total deaths are 206,482. It caused global economic and public health emergencies. It spreads by direct or indirect contact with a respiratory droplets from infected individuals(Holshue et al. 2020). Several steps like physical distancing, quarantine, and other sanitizing habits have shown some success in slowing down the pandemic but it is still far from being contained in most countries.

There is no known treatment or vaccine available. The antimalarial drug chloroquine and antibiotic azithromycin have showed some hope against COVID-19 but its efficacy has been recently debated (Syal 2020). Other potential therapeutic agents tried so far include Remdesivir, Lopinavir / Ritonavir (Kaletra), Tocilizumab(Actemra), Remdesivir have shown some promise in controlling the COVID-19 disease. (Holshue et al. 2020; Lu 2020; Russell et al. 2020; Wang et al. 2020). Most people in developing countries cannot afford costly therapeutic interventions like mechanical ventilators and prolonged lockdown, pandemic must be curtailed so that health infrastructure can manage it efficiently. As there is no known treatment and vaccine available, herd immunity can help in decreasing its spread. (Syal 2020)

Immunotherapy and plasma therapy has been used effectively as a therapeutic option against many viral infections. The main methods in immunotherapy include several vaccines and monoclonal antibody candidates. Convalescent plasma (CP) therapy has been used for the prevention and treatment of many infectious diseases for more than one century. In both SARS-CoV (Severe Acute Respiratory Syndrome Corona Virus), and SARS-CoV-2 viruses’ entry into the host cells is mediated by the interaction of the receptor-binding domain (RBD) in S protein on virus outer-membrane and angiotensin-converting enzyme 2 (ACE2) on cell. So, these proteins can be the major potential targets for immunotherapy (AminJafari and Ghasemi 2020; Duan et al. 2020b). CP refers to plasma that is collected from individuals, following the resolution of infection and development of antibodies. Over the past two decades, CP therapy was successfully used in the treatment of SARS MERS (Middle East Respiratory Syndrome), and 2009 H1N1 pandemic with satisfactory efficacy and safety (Cheng et al. 2005; Zhou et al. 2007; Hung et al. 2011; Ko et al. 2018).

A meta-analysis from 32 studies of SARS coronavirus infection and severe influenza showed a statistically significant reduction in the pooled odds of mortality following CP therapy, compared with placebo or no therapy (odds ratio, 0.25; 95% confidence interval, 0.14-0.45) (Mair-Jenkins et al. 2015). Since SARS, MERS, and COVID-19 shares similar virologic and clinical symptomatology (Lee and Hsueh 2020), CP therapy might be a promising treatment option for COVID-19 treatment (Chen et al. 2020). Studies done by Cheng (Cheng et al. 2005) in Hong Kong reported that in 2003 during SARS outbreak, patients who received convalescent plasma had a lower mortality rate (12.5%) compared with the overall SARS-related mortality for admitted patients (n = 299 [17%]).

Convalescent plasma (CP) has also been used in the COVID-19 pandemic; limited data from China suggest clinical benefit, radiological resolution, reduction in viral loads, and improved survival(Bloch et al. 2020). While fractionated plasma products (e.g. hyperimmune globulin, monoclonal antibodies) and/or vaccination may offer durable therapeutic options, human anti-SARS-CoV-2 plasma is the only therapeutic option that is immediately available for use to treat COVID-19(Bloch et al. 2020). Studies done by Shen et al, and Duan et al (Duan et al. 2020b; Shen et al. 2020) concluded that convalescent plasma therapy is well tolerated and could potentially improve the clinical outcome. They collected CP from patients three weeks after they recovered from COVID-19, and from the patients who were having normal body temperature for > 3 days, not having respiratory symptoms, and there two consecutive PCR SARS COV-2 test 24 hours apart has been negative. Their study result showed a decline in inflammatory markers, improvement in patient’s antibody titer and PCR SARS COV-2 became negative. CFR (Case Fatality Rate) was 0 in the study done by Duan et al. However there were several limitations of the study including small sample size, lack of randomized double-blind controlled study, those patients received other medications like antivirals (Kaletra), steroids at the same time, and it is difficult to attribute all improvement to plasma therapy alone. Their study indicates that convalescent plasma therapy could be the most critical weapon in the fight with COVID-19 in severe cases. Survivors of the COVID-19 may play a key role in both herd immunity as well as the availability of plasma therapy. (Syal 2020).

Since the effective vaccine and specific antiviral medicines are unavailable, there is an urgent need to look for an alternative strategy for COVID-19 treatment, especially among severe patients. Our aim for the present study is to create a compartmental mathematical model for COVID-19 transmission and then apply various control parameters like self-isolation, quarantine of infected individual and hospitalization to receive medication (immunotherapy and convalescent plasma therapy) and see their effects on recovery from COVID-19 disease.

## 2. Mathematical model

In this section, a basic model for COVID-19 transmission dynamics among humans is constructed. Infected individuals who regularly come in contact with exposed class, for example, vegetable vendor, grocery store-keeper, policeman or security man, delivery man etc., they all unknowingly spreads infection at high rate. Hence, these population class are considered to be a super active spreader of the infection. Remaining all infected (symptomatically and asymptomatically) individuals who are capable to infect others are considered to be an active spreader of COVID-19 infection. The model contains these two different class of infected classes who are the spreader of the infection and accelerate the intensity of the COVID-19 outbreak. In some cases strong immunity of infected individual can defeat the infection and can make individual disease free without hospitalisation. Moreover, in some cases recovered individuals again shows symptoms of the disease and hence they need to get hospitalisation again. These two situation are also considered in the present model.

In the model, total population is divided in seven compartments: exposed (not infectious) individuals (*E*), infected individuals (*I*), infected individuals who are active spreader (*A*), infected individuals who are super active spreader (*S*), quarantined individuals (*Q*), hospitalised individuals (*H*) and recovered individuals (*R*); Fig. 1. Using this model, formulated dynamics of non-linear differential equation is shown below.

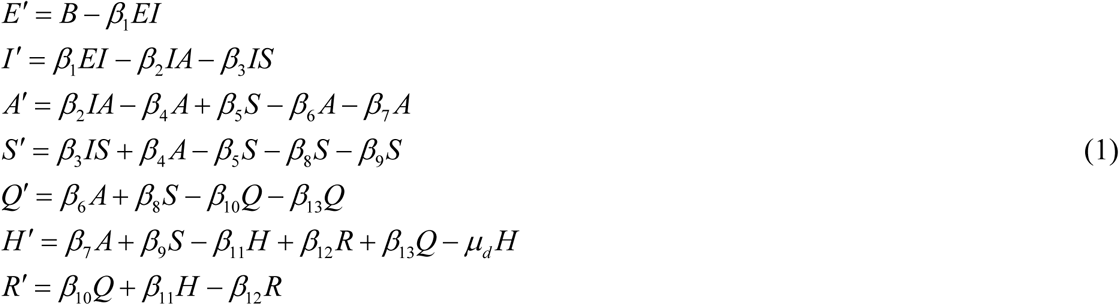

**Figure 1.**
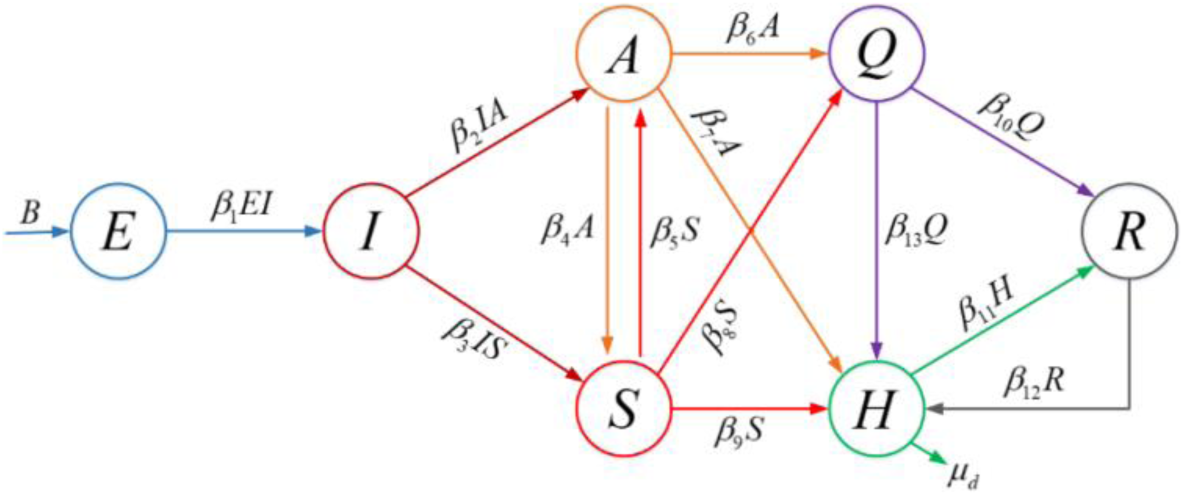
System diagram of COVID-19 transmission model

Parameters used in the model are listed in the table 1.

**Table 1.** Parameters used in the model. Note that, data for total number of infected cases, critical cases, hospitalised cases and deaths due to COVID-19 are taken from worldometers on 25th April, 2020, (Worldometer 2020). Approximate parametric values are calculated using the data available and some are assumed.

| Parameters |  | Parametric value |
| --- | --- | --- |
| $B$ | Recruitment rate of exposed individuals | 0.6 |
| $\beta_1$ | Recruitment rate of infected individuals | 0.17 |
| $\beta_2$ | Rate at which infected individuals become active spreader | 0.40 |
| $\beta_3$ | Rate at which infected individuals become super active spreader | 0.20 |
| $\beta_4$ | Rate at which active spreader become super active spreader | 0.05 |
| $\beta_5$ | Rate at which super active spreader become active spreader | 0.30 |
| $\beta_6$ | Rate at which active spreader moves to quarantine | 0.02 |
| $\beta_7$ | Rate at which active spreader moves to hospitalisation | 0.70 |
| $\beta_8$ | Rate at which super active spreader moves to quarantine | 0.30 |
| $\beta_9$ | Rate at which super active spreader moves to hospitalisation | 0.70 |
| $\beta_{10}$ | Recovery rate of quarantined individuals without hospitalisation | 0.07 |
| $\beta_{11}$ | Recovery rate of hospitalised individuals | 0.44 |
| $\beta_{12}$ | Rate at which recovered individuals again moves to hospitalisation | 0.30 |
| $\beta_{13}$ | Rate at which quarantined individuals get hospitalisation | 0.60 |
| $\mu_d$ | Mortality rate of COVID-19 | 0.1077 |

### 2.1. Well-posedness of the solution

In this sub-section, we observe that the solutions of the system (1) are non-negative and bounded if initial conditions are non-negative. Since the model contains only human population, only non-negative initial conditions are used. Also all the parameters used in the model are considered non-negative. We know that (*E*(*t*), *I*(*t*), *A*(*t*), *S*(*t*), *Q*(*t*), *H*(*t*), *R*(*t*)) ≥ 0 if (*E*(0), *I*(0), *A*(0), *S*(0), *Q*(0), *H*(0), *R*(0)) ≥ 0. Based on system (1), we have *N = B* − *μ_d_H*. Where *N* = *E + I + A + S + Q+H + R*. When *t* → *∞*, we have *N ≤ B*, since *μ_d_* ≥ 0. Hence *N* is bounded and the feasible region is Λ:

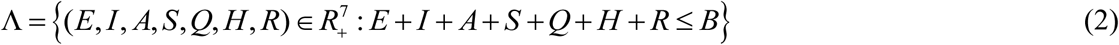

### 2.2. Basic reproduction number

The basic reproduction number *R*_0_ is defined as the average number of secondary infected cases rising from an average primary case in an entirely exposed/susceptible population.

The solution of the system (1), the endemic equilibrium point 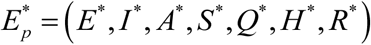 is as follow:

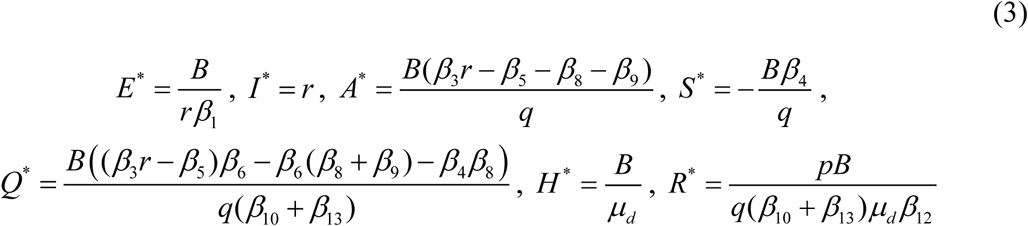

where,

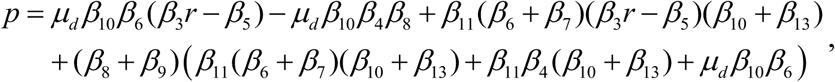

*q* = (*β*_6_ + *β*_7_)(*β*_3_*r* − *β*_5_)−(*β*_8_ + *β*_9_)(*β*_4_ + *β*_6_ + *β*_7_) and *r* is the highest root of a polynomial *m*(*z*) = *a*_0_*z*^2^ + *a*_1_*z* + *a*_2_ = 0, coefficients of the polynomial are: *a*_0_ = *β*_2_*β*_3_, *a*_1_ = (*β*_2_ (*β*_5_ + *β*_8_ + *β*_9_) + *β*_3_ (*β*_4_ + *β*_6_ + *β*_7_)) and *a*_2_ = (*β*_5_ (*β*_6_ + *β*_7_) + (*β*_8_ + *β*_9_)(*β*_4_ + *β*_6_ + *β*_7_). This *R*_0_ based on endemic equilibrium point can be calculated using next generation matrix method(Diekmann et al. 1990; Garba et al. 2008). The above system (1) can be written in the following form using matrices.

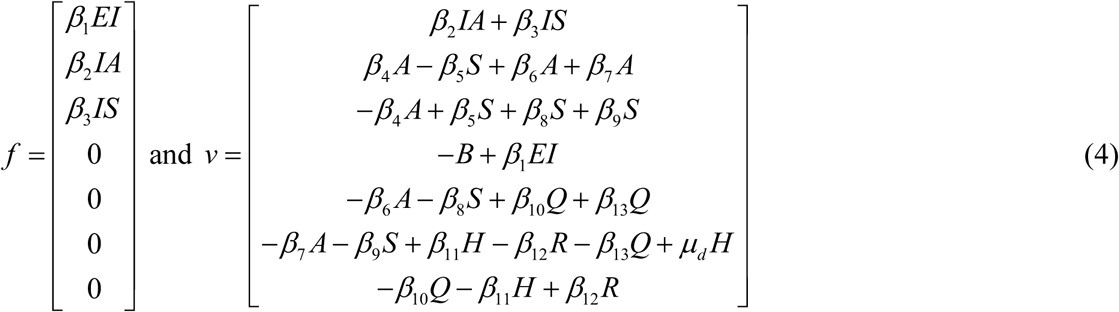

Note that, matrix *F* shows the new infectious rates and matrix *V* shows other rates transferred in between the compartments, are given respectively by:

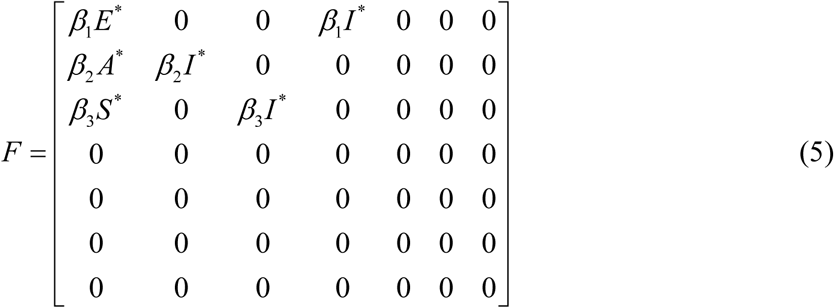

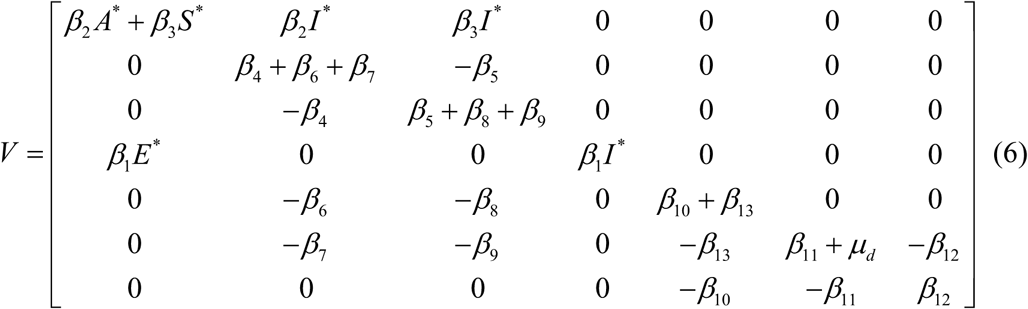

Hence, the basic reproduction number (*R*_0_), is given by

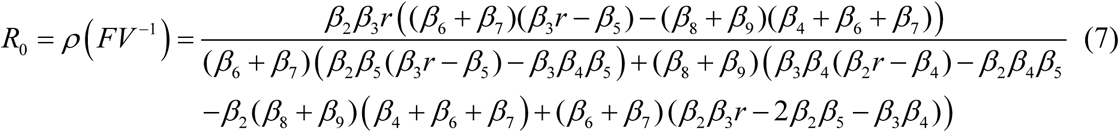

where *ρ* is the dominant eigenvalue in magnitude of the matrix *FV*^−1^. After substituting all parametric values from table 1, we get the threshold value *R*_0_ =3.4709 which represent the average number of secondary cases generated by an infected individual in completely susceptible population. From this calculation, we can say the model or the current outbreak is in highly unstable, hence, certain control strategies are very essential to impose to come out from pandemic situation.

## 3. Optimal control

In this section, we extend the system (1) to include five time dependent control strategies, *u*_1_(*t*), *u*_2_(*t*), *u*_3_(*t*), *u*_4_(*t*) and *u*_5_(*t*), regarding isolation of infected individuals and medication to improve immunity to fight against the COVID-19 outbreak.

Here, control variables *u*_1_(*t*) and *u*_2_(*t*) measures the quarantine or isolation of the individuals who are spreader and super spreader respectively. We can provide herd immunity or herd protection to those who are not immune to the COVID-19 disease by improving immunity in most of the population. Also strong immunity of large mass can control further spread of COVID-19 infection. The control variable *u*_3_(*t*) indicates the strategy which increases the herd immunity which helps quarantined individuals to get recovered without medication. The control variable *u*_4_(*t*) suggests to provide proper immunotherapy (include several types of vaccines, monoclonal antibody candidates, and etc.) to hospitalised individuals which helps them to fight against the viral infection. In present situation, where proper vaccination is not available for the COVID-19, a convalescent plasma therapy have sparked a ray of hope. The Convalescent plasma therapy’s effects as treatment for Covid-19 has been tested positive with no severe adverse effects (Duan et al. 2020a, 2020b; Shen et al. 2020). The control variable *u*_5_(*t*) is used to support convalescent plasma therapy to improve immunity of critically infected individuals. However, the limitation of this control strategy is, if plasma therapy is not used properly and carefully, it can create more serious complications.

Under these assumptions, the COVID-19 model (1) is re-constructed by including control variables on Fig. 2:

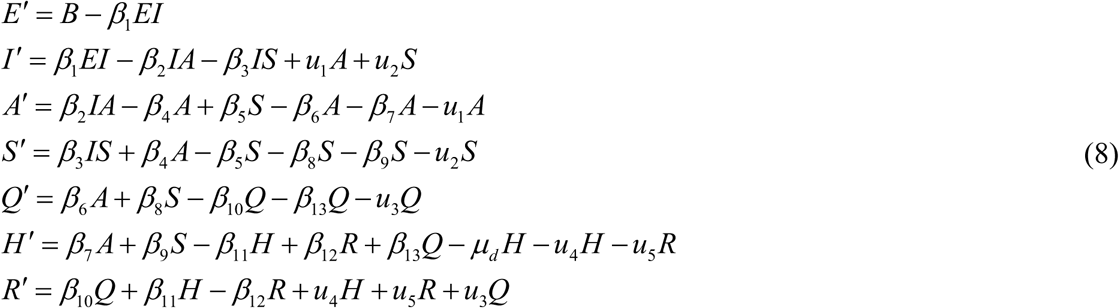

**Figure 2.**
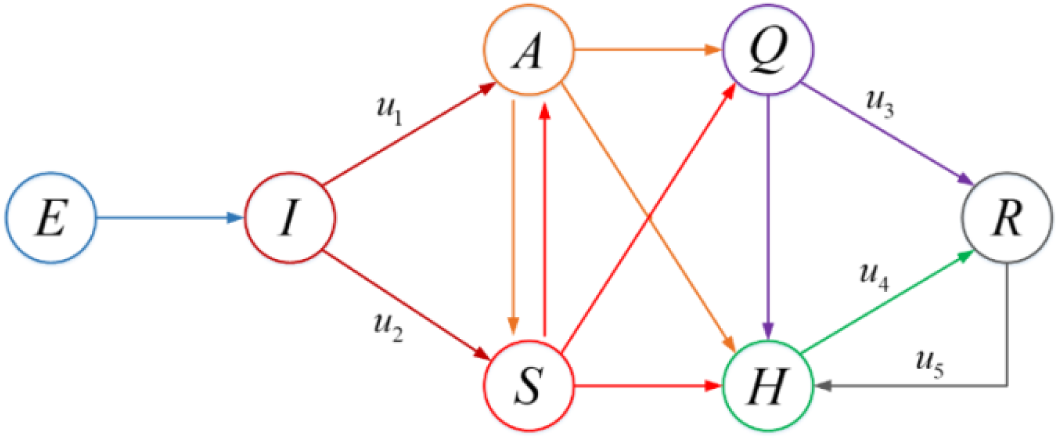
COVID-19 model with control variables

According to this extended model, the optimal control problem with the objective function is formulated by

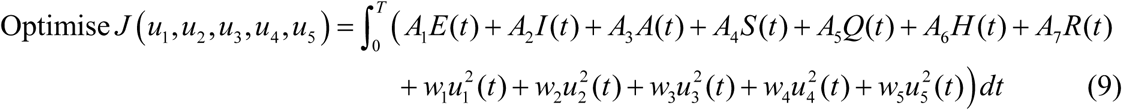

The objective is to minimise active and super active spreaders, increase recovery of hospitalised and quarantined individuals. In equation (9), *A_i_*, *i* = 1,2,…7, are weight constants of the state variables and *w_j_*, *j* = 1,2,…,5 are weight constants of respective control variables. Our goal is to determine optimal control functions 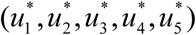, such that

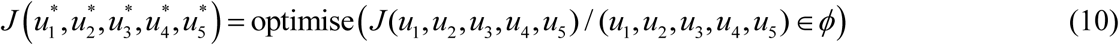

subject to the modified system (8), where *ϕ* is a control strategy set.

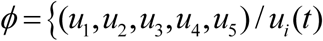 is Lebesgue measurable on[0,T],0 ≤ *u_i_*(*t*) ≤ 1, *i* = 1,2,…,5}

### Theorem 1

Consider the objective function (9) with (*u*_1_, *u*_2_, *u*_3_, *u*_4_, *u*_5_) ∈ Γ subject to the constraint state system (8) then there exist 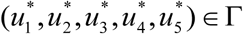 such that 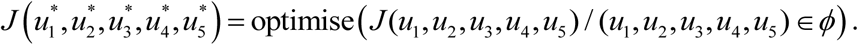.

#### Proof

The integrand, 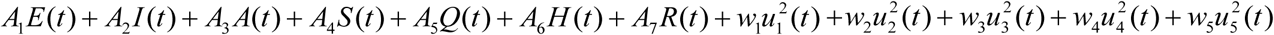 of the objective function (9) is convex in the set *ϕ*. The control strategy set *ϕ* is also close and convex by definition. Since the model (8) is bounded and linear in the control variables, the conditions for the existence of optimal control are satisfied (Fleming and Rishel 1975).

### 3.1 Optimality system

Let us convert the problem into a problem of maximizing a Lagrangian function *L*, with respect to all control variables *u*_1_, *u*_2_, *u*_3_, *u*_4_, and *u*_5_. For necessary condition of an optimal control problem, Pontryagins maximum principle (Pontryagin 2018)is used.

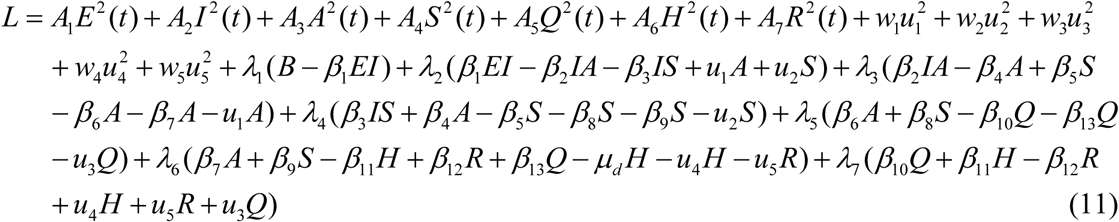

For given an optimal control 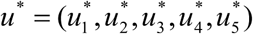 and corresponding state solutions of the system (8), there exist adjoint functions, 4, *i* = 1,2,…7, which are

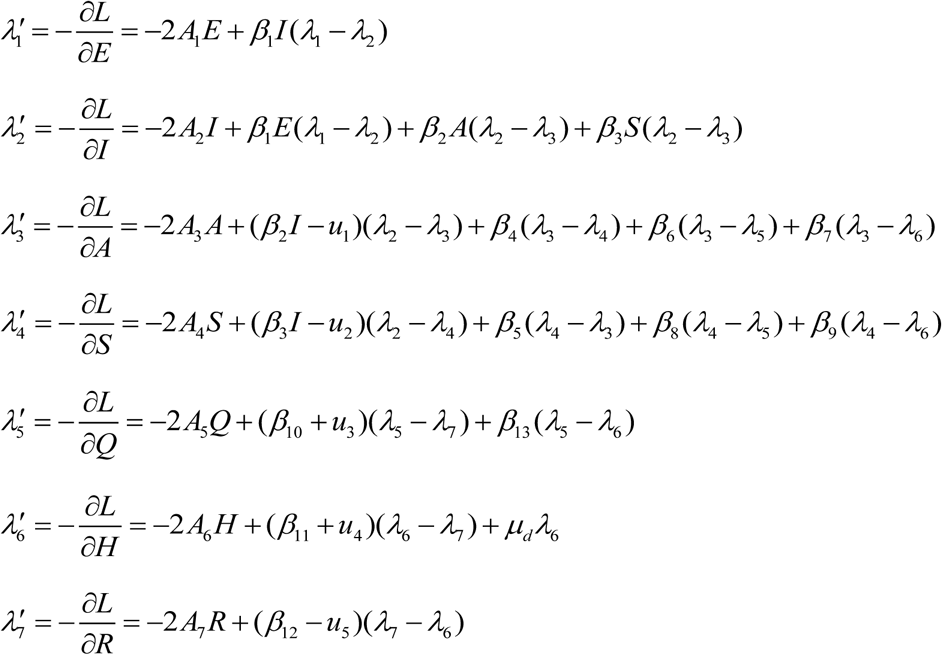

The terminal conditions are *λ_i_* (*T*) = 0, for *i* = 1,2,…7. The optimal control variables 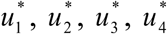 and 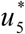 can be solves using optimality conditions

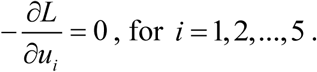

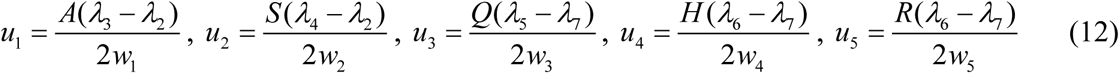

Moreover, optimal control strategies 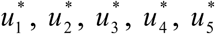 are given by:

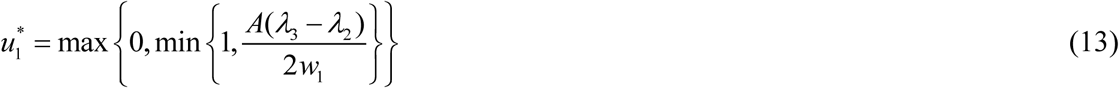

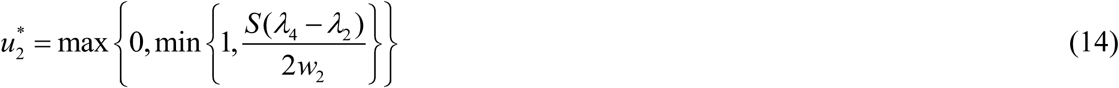

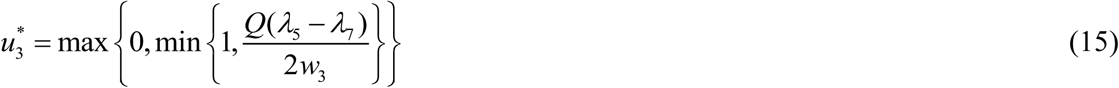

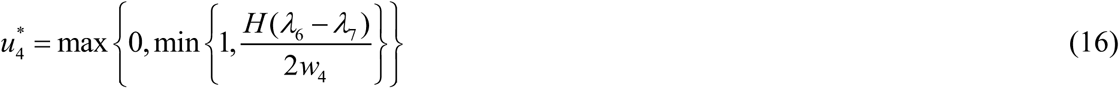

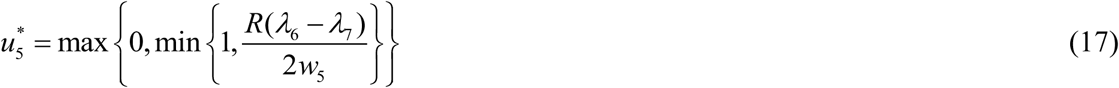

## 4. Numerical simulation

This section attempts to describe the graphical representation of variations in the model under influence of optimal control strategies. The initialisation of exposed, infected, active and super active spreader, quarantined, hospitalised and recovered is given by *E*(0) = 10, *I*(0) = 8, *A*(0) = 4, *S* (0) = 4, *Q*(0) = 3, *H* (0) = 4 and *R*(0) = 2 respectively.

Figure 3 shows variation with time in each compartment of the model. In the initial week of the outbreak, higher intensity of infected, active and super active spreader is observed, after that, they decreases with time and become negligible in 5-6 weeks. In this duration, since individuals in exposed and hospitalised class are still present, again growth in infection is observed after 7-8 weeks of the outbreak. From this graph, we can say that without any control strategies the infection can re-emerge in the society after some time of duration. Hence proper control strategies are essential to break the periodic chain of this infection.

**Figure 3.**
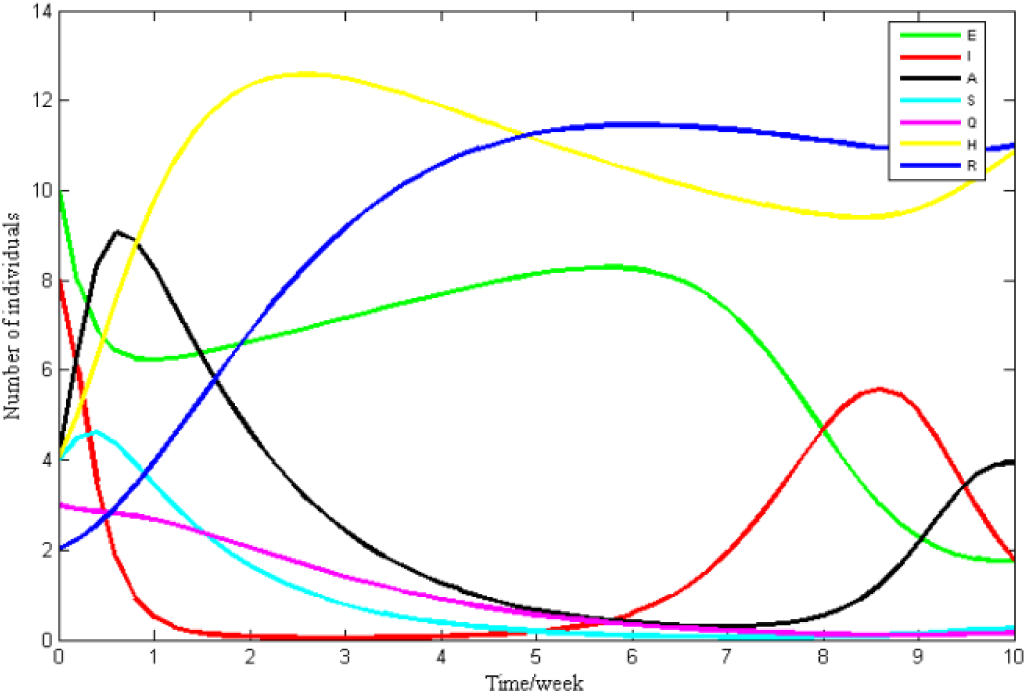
Variation in compartment with time

Figure 4(a) and 4(b) displays the effect of super active spreaders on class of exposed and infected individuals respectively. Figure 4(a) shows that exposed individuals are getting infected by super active spreaders at high rate. Figure 4(b) shows that infected individuals are moves towards the super active spreaders and they have tendency to becoming a super active spreader, while super active spreaders are also moving towards the infected class at lower intensity. This indicates that many super active spreaders getting aware of the disease transmission and they stop spreading the infection by isolation.

**Figure 4.**
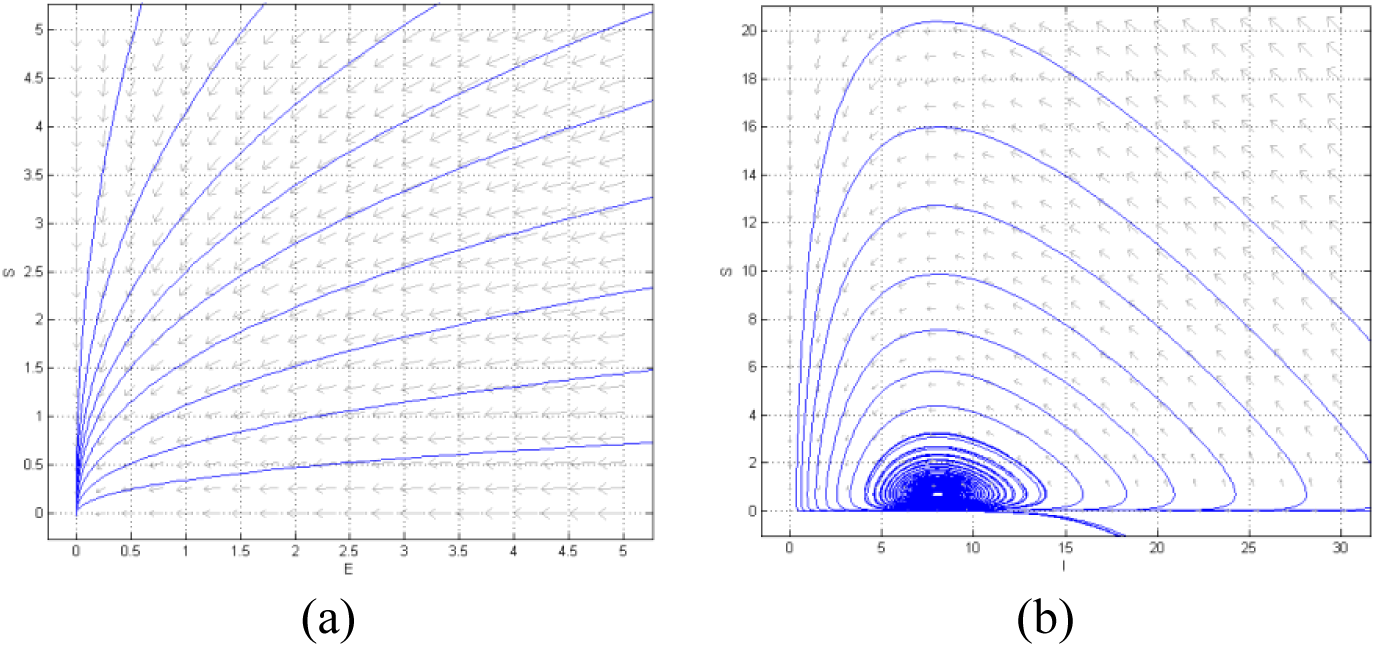
Effect of super active spreader on transmission of COVID-19

Figure 5(a) shows the intensity of super active spreaders moves towards hospitalisation and figure 5(b) shows the recovery frequency of super active spreaders.

**Figure 5.**
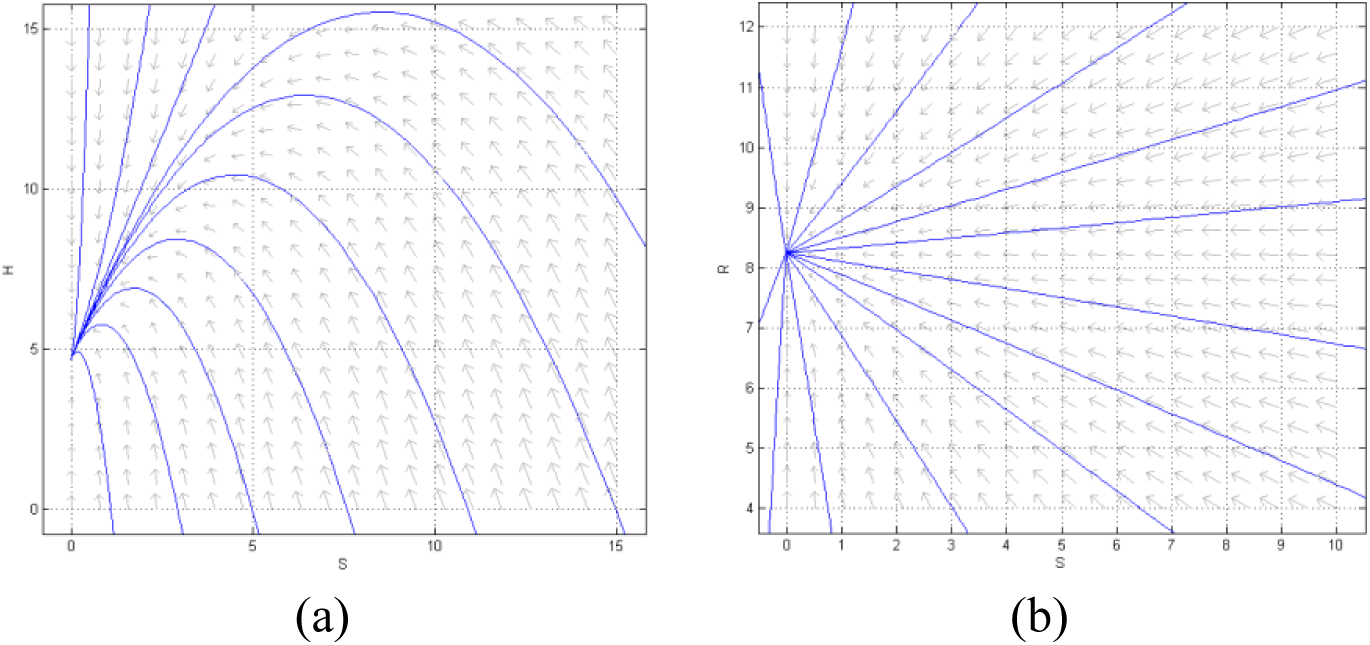
Controlling super active spreaders of COVID-19

Periodic transmission of infection through the compartments with respect to time (in week) is observed in the figure 6. Figure 6(a) and 6(b) shows periodic transmission of active and super active spreaders respectively, though hospitalisation and recovery class. This scenario suggest that after recovery, still there is a chance of infection. Figure 6(c) and 6(d) shows the intensity and periodicity of infected individuals getting recovered by their strong immunity after self-quarantine. Figure 6(e) shows periodic oscillations between infected, hospitalised and recovered class. It can be observed from figure 6(f) that the infection is moving periodically around the class of active spreaders, hence we can say that, largest persisting period of the infection is when it is in class of active spreaders.

**Figure 6.**
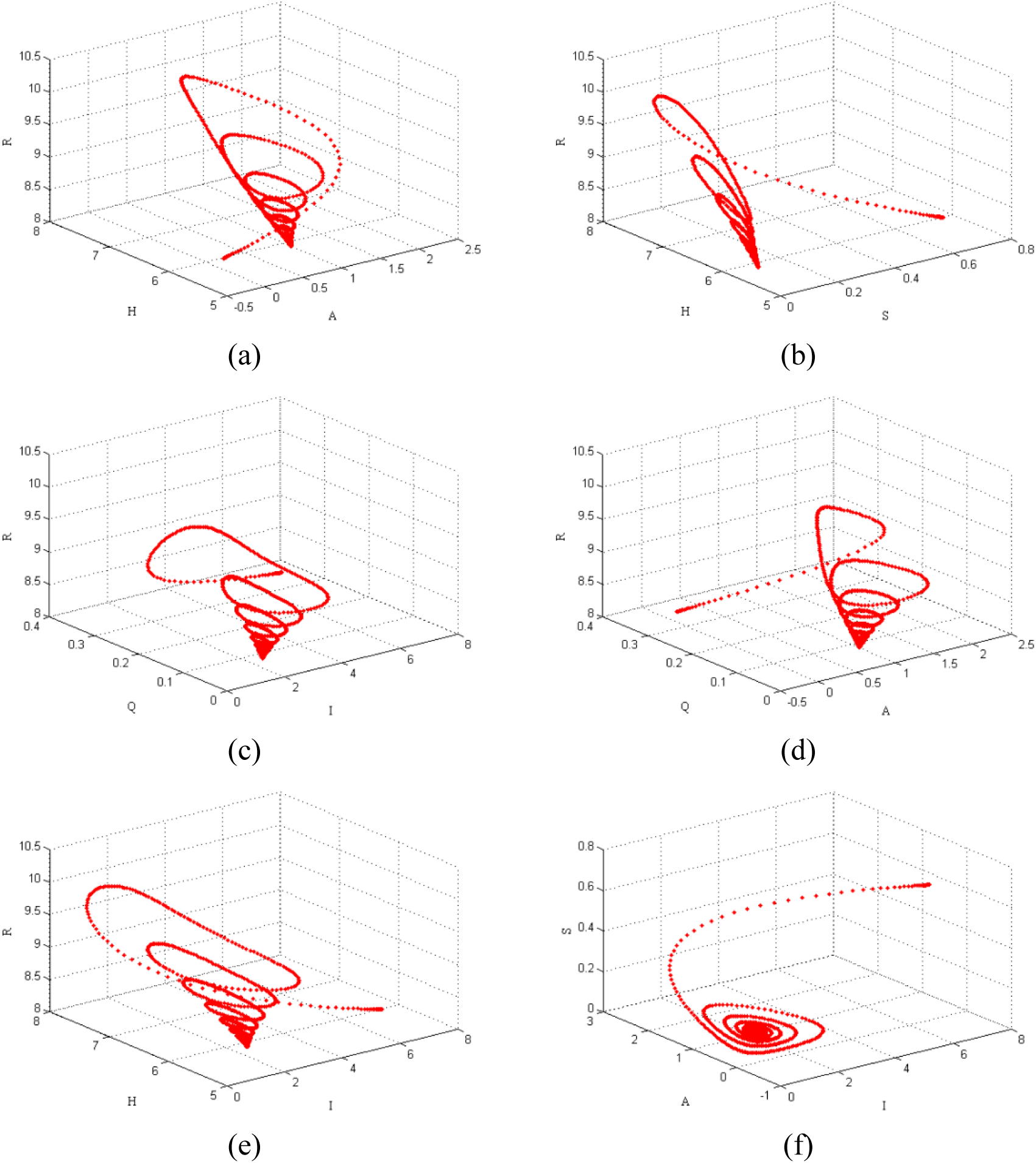
Phase portrait diagram of COVID-19 model

Figure 7 shows the oscillations in the model compartments during the outbreak. In the initial days of the outbreak, noteworthy oscillations in the model are observed, moreover the figure show that after 100-120 days, the model shows its asymptomatic stability.

**Figure 7.**
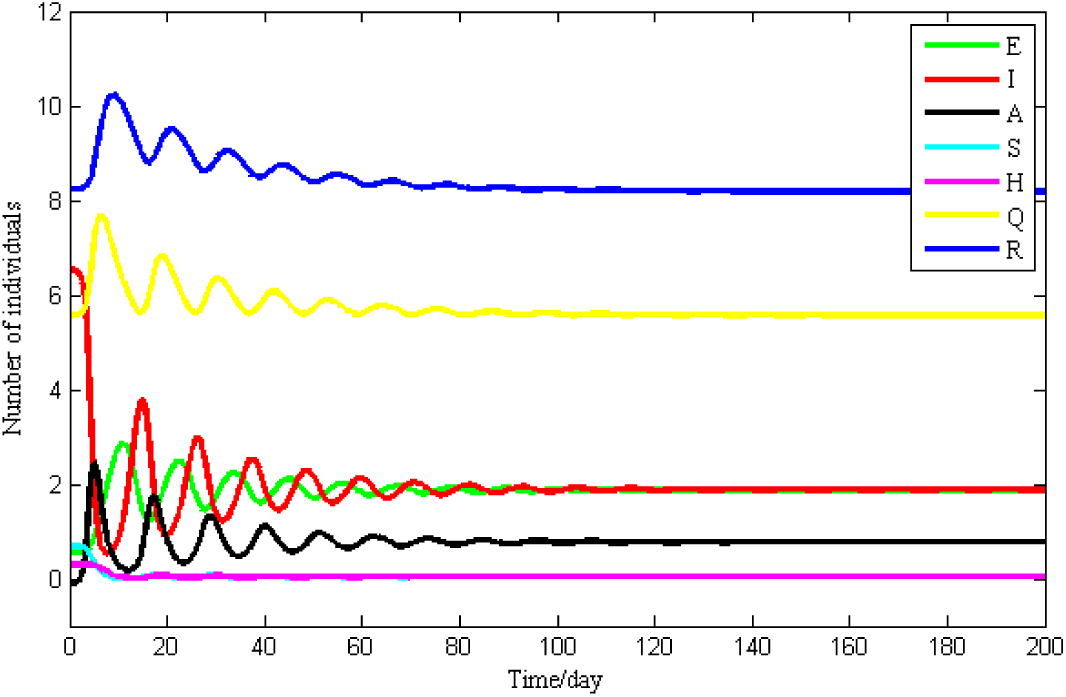
Oscillations in the model during the COVID-19 outbreak

### 4.1. Simulation influenced by optimal control theory

In this section, effect of all control strategies on transmission of COVID-19 is observed graphically.

Figure 8 shows variation in each compartment under influence of with and without control strategies. It is observed that COVID-19 outbreak can be controlled up to significant level in three weeks after applying all the control strategies together. Super active spreaders are the major threatening problem during this pandemic outbreak. Figure 8(d) shows that the intensity of super active spreader is controlled notably after applying the controls which is major factor to reduce spread of COVID-19. Under this controlled situation reduction in hospitalisation and raise in recovery from infection is clearly observed in figure 8(f) and 8(g) respectively.

**Figure 8.**
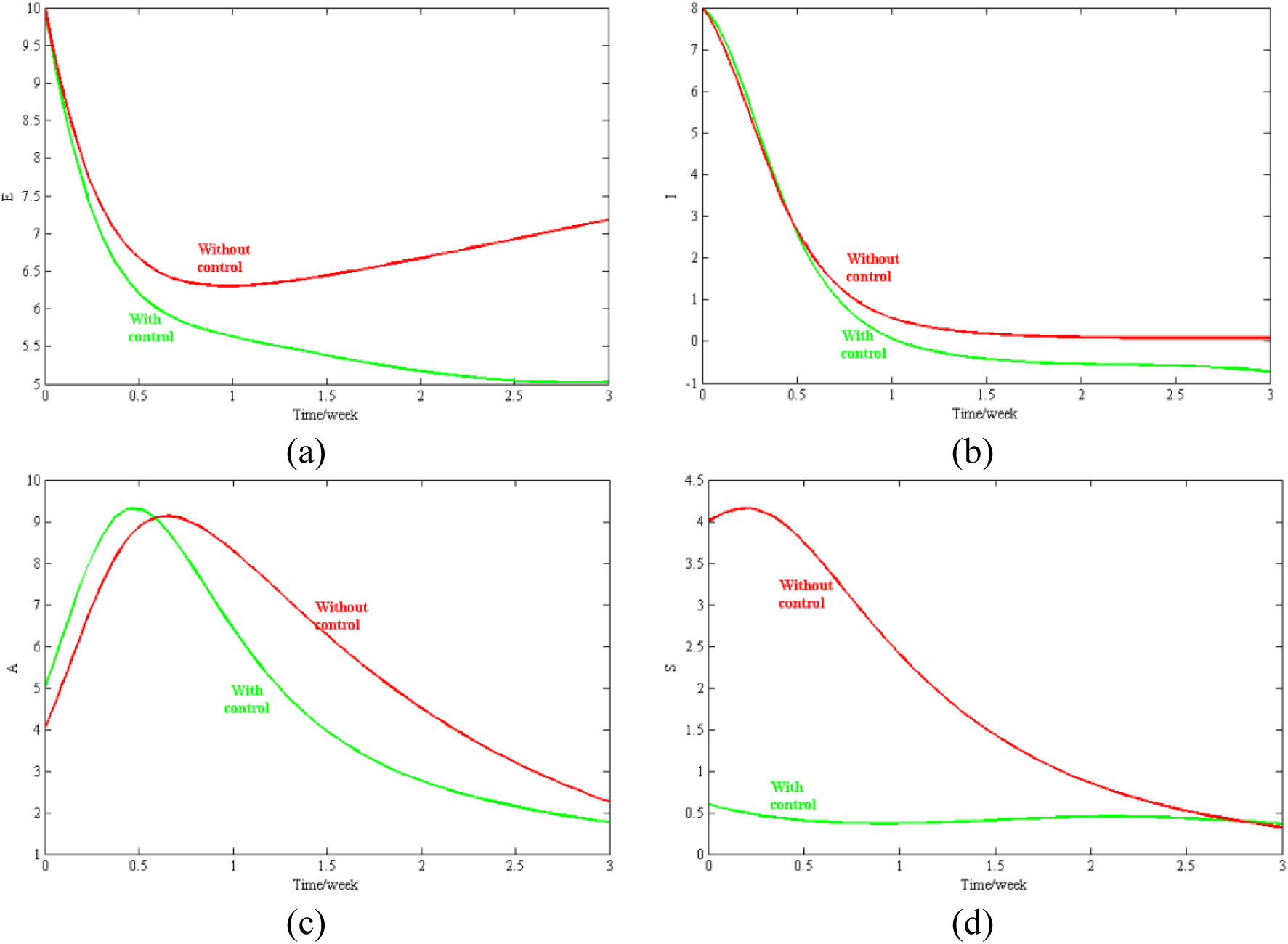
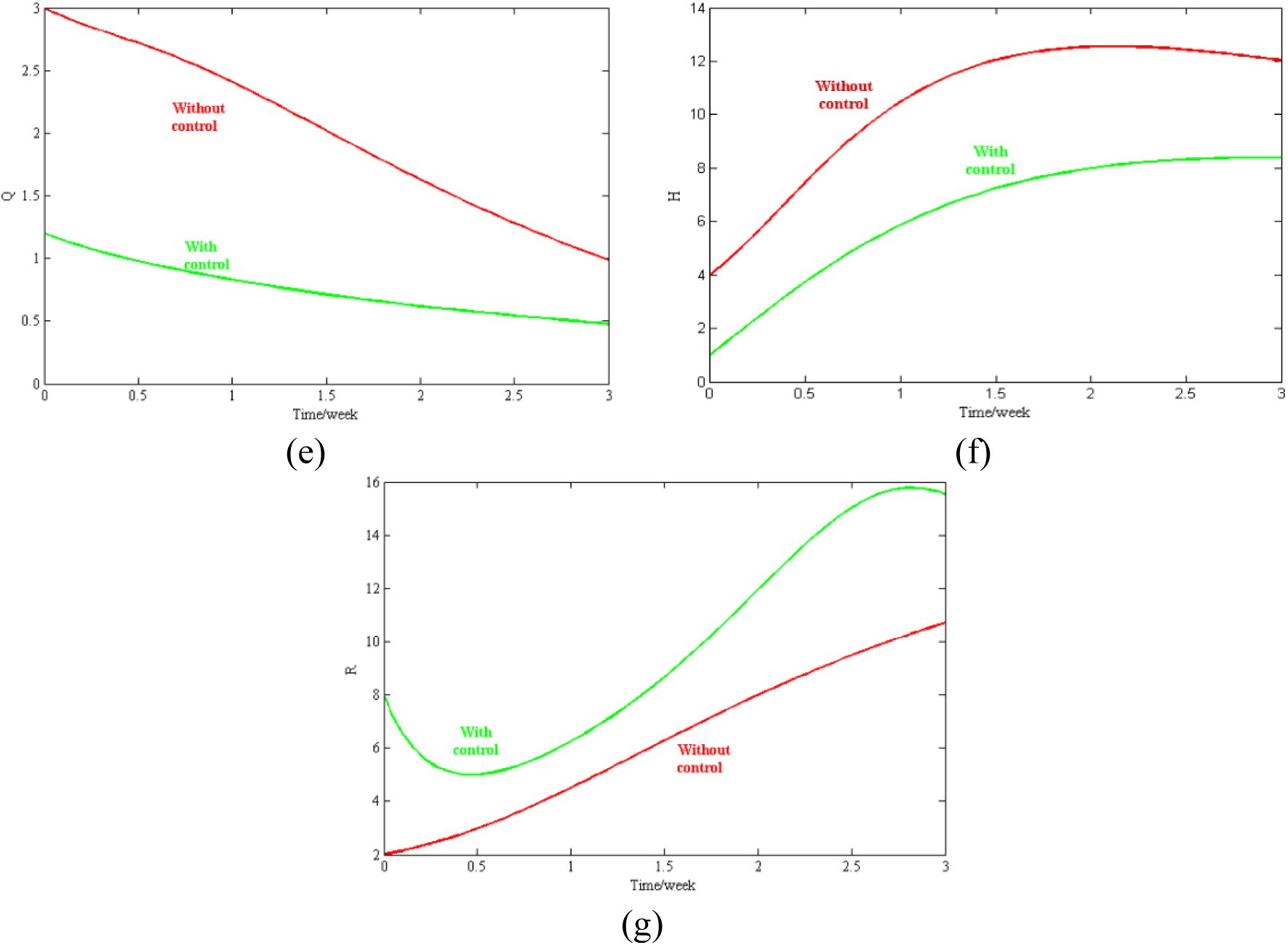
Change in each compartment with and without controls

Figure 9 shows an individual effect of immunotherapy and plasma therapy on class of recovered individuals. The figure indicate that, initially immunotherapy is highly effective on hospitalised infected individuals, moreover in long term, better results are observe on recovered class when plasma therapy is applied.

**Figure 9.**
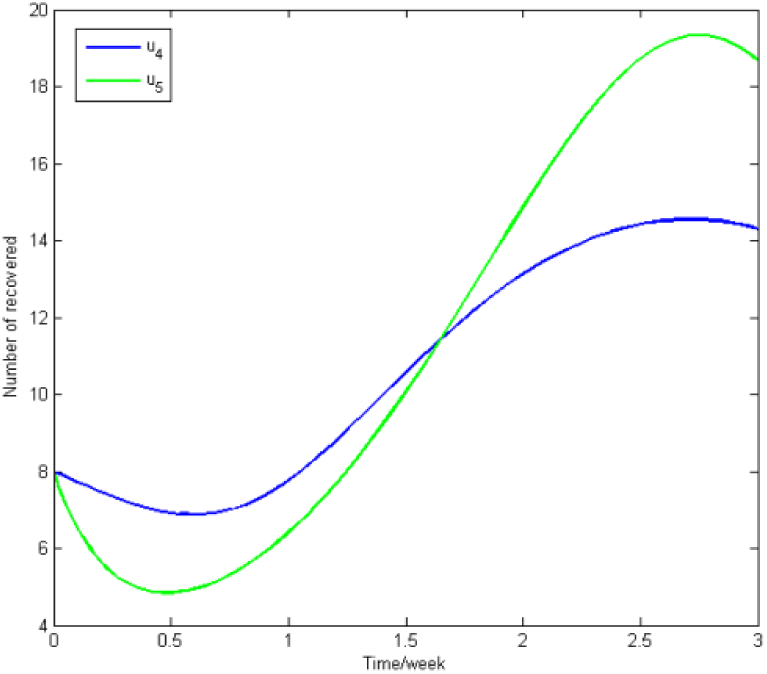
Effect of immunotherapy and plasma therapy on recovered class

N.B. On y-axis reading is x*10.

Deviation in the intensity of control strategies with time is shown in figure 10. Isolation varies from 10% to 32%, maximum of 21% quarantine facilities should be used. Using preventive measures, herd immunity can be attained to be 20%. 33% immune therapy and 10% plasma therapy should be applied together to fight back COVID-19 outbreak in around 50 days.

**Figure 10.**
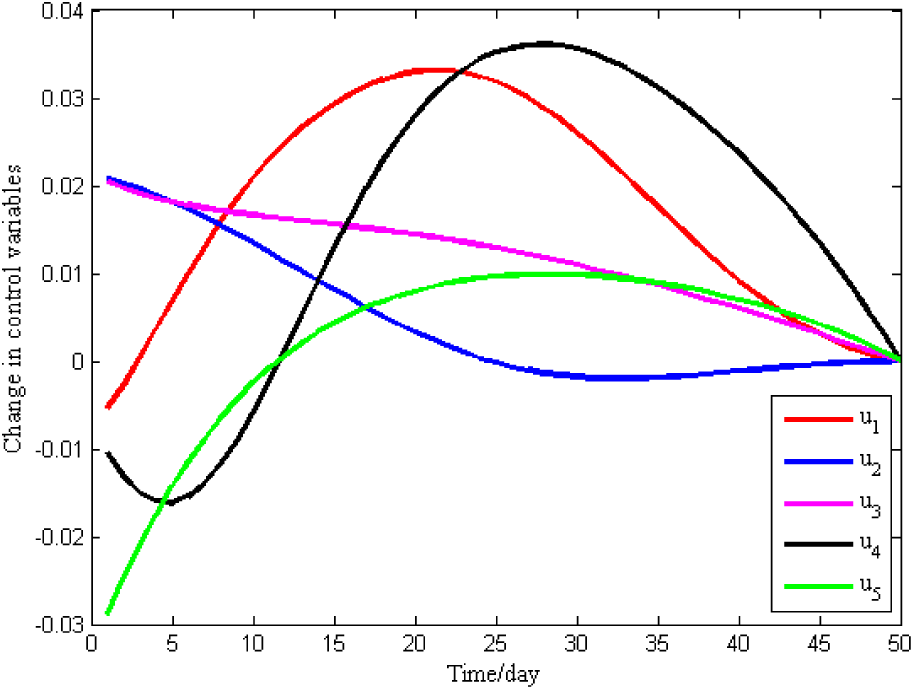
Change in control variables with time

### 4.2. Bifurcation analysis

In the current section, backward bifurcation theory is analyse to understand the behaviour of threshold value of the COVID-19 model. Note that, non-negative equilibria (3) of the COVID-19 model satisfies the quadratic in the infected class (*E*). Positive equilibrium of the system is achieved by solving the quadratic equation for (*z*). The bifurcation analysis helps to validate the qualitative information about the basic reproduction number.

The bifurcation diagram is shown in figure 11, where blue vertical line indicates the value of the critical point *R_c_*, which is 2.23. We can say that this is the point from which system’s stability switches from unstable to stable state. To effectively control the spread of COVID-19, the basic reproduction number should be brought below *R_c_*. Red vertical line in the figure 11 shows the numerical value of basic reproduction number. By observing current situation of this pandemic outbreak, it is very hard to bring the value of *R*_0_ below *R_c_* in short time period.

**Figure 11.**
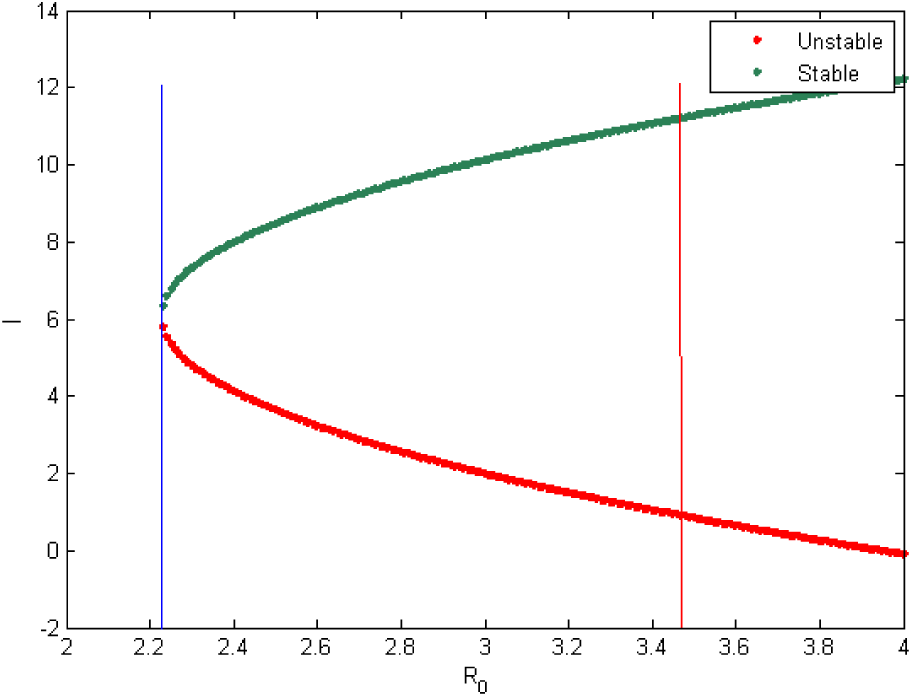
Bifurcation diagram for the COVID-19 model

Figure 12 represents the bifurcation diagram of the model with respect to the rate at which infected individuals become super active spreader (*β*_3_). Here, the maximum and minimum values of the fluctuations are plotted in blue and red colours respectively. Since the super spreader can create more infection, notable changes in the recovery rate is observed when a gradual change made in the parameter *β*_3_, but it will reduce later.

**Figure 12.**
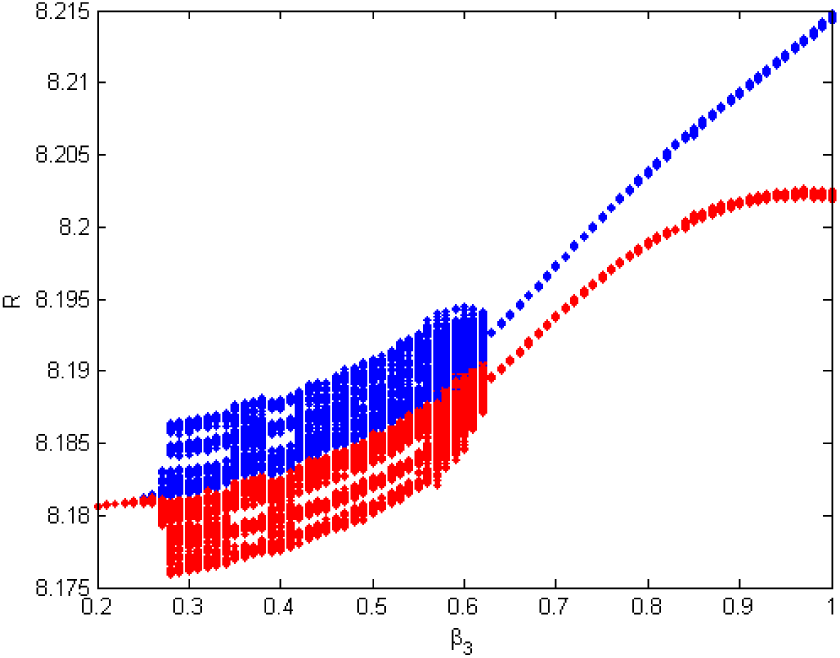
Bifurcation w.r.t *β*_2_

## 5. Discussion and Conclusion

Our study showed that in the initial week of the outbreak, there is a large number of people who are infected, and behave as active and super active spreader but in about 5-6 weeks of the time they become negligible. Many super active spreaders are becoming aware of the mode of disease transmission and stop spreading the infection by self-isolation. However, there are still exposed and hospitalized individuals who are responsible for resurgence or second wave of infection at around 7-8 weeks of time.

Bifurcation analysis showed that R_c_ (critical point) is 2.23 and R_o_ is 3.5, and by observing the current situation of this pandemic, it is very difficult to bring the value of R_o_ below R_c_ in a short period of time. All the control measures including self-isolation, quarantine, herd immunity, immunotherapy, and plasma therapy should be applied together to fight back COVID-19 in around 50 days. Effectiveness of self-isolation varies between 10-32%, quarantine is about 21%, herd immunity is about 20%, immune therapy is 33% and plasma therapy is about 10%. Optimal timing of the plasma therapy is around 15 days from the infection while immunotherapy should be implemented earlier to get maximum benefit.

Thus we conclude that the COVID-19 outbreak can be controlled up to a significant level three weeks after applying all the control strategies including self-isolation, quarantine, and hospitalization together. Furthermore, proper control strategies are of paramount importance in breaking this periodic chain of infection and preventing the resurgence of infection. Super active spreaders are the major threatening problem during this pandemic outbreak. Our results show that the super active spreaders can be controlled notably by optimal control strategies. These strategies lead to a reduction in hospitalization and a rise in recovery from infection. Immunotherapy is highly effective initially in hospitalized infected individuals however better results were seen in the long term with plasma therapy.

We also suggest certain policies to make sure that plasma therapy is available on a larger scale. The Physician can motivate patients at the time of discharge to donate plasma in the near future (once they are eligible to donate). Furthermore, the government should expand plasma collection capabilities. They need to create infrastructure and remove certain barriers for agencies in order to prioritize collecting plasma and making it available on a larger scale for treatment at subsequent waves of COVID-19 infection.

Preliminary data using immunotherapy and plasma therapy against the rapidly increasing number of COVID-19 cases provides an unprecedented opportunity to perform a large-scale randomized clinical trial, to study the efficacy of this treatment against a viral agent. If the results of rigorously conducted investigations demonstrate consistent efficacy, the use of these therapies could help change the course of this pandemic.

## Data Availability

Our data derived from our mathematical calculations in the paper that are available to all the readers.

## Acknowledgement

All the authors are thankful to DST-FIST file # MSI-097 for technical support to the Department of Mathematics, Gujarat University. The second author (AHS) is funded by a Junior Research Fellowship from the Council of Scientific & Industrial Research (file no.-09/070(0061)/2019-EMR-I). The third author (ENJ) is funded by UGC granted National Fellowship for Other Backward Classes (NFO-2018-19-OBC-GUJ-71790).

